# Neuroimaging and Cognitive Testing in Healthy Aging Adults using a Portable Low-Field MRI Scanner and Web-Based Assessment

**DOI:** 10.1101/2022.08.22.22278933

**Authors:** Sean CL Deoni, Phoebe Burton, Jennifer Beauchemin, Rosa Cano-Lorente, Matthew D. De Both, Megan Johnson, Lee Ryan, Matthew J. Huentelman

## Abstract

Consumer wearables and health monitors, internet-based health and cognitive assessments, and at-home biosample (e.g., saliva and capillary blood) collection kits are increasingly used by public health researchers to recruit and follow large study populations without requiring intensive in-person study visits. In addition to reducing participant time and travel burden, remote and virtual data collection allows individuals who live long distances from a hospital or university research center, have limited time or mobility, or who lack access to transportation to participate. Unfortunately, studies that include magnetic resonance neuroimaging can be particularly burdensome given the infrastructure requirements of 1.5, 3, and 7 Tesla scanners. As a result, they often omit socially, economically, and educationally disadvantaged individuals. Portable lower magnetic field strength systems offer the potential to perform neuroimaging at a participant’s home and convenience. In this work, we present the first report of associations between brain morphometry and cognitive performance assessed using a portable low-field MRI “scan van” and an established online assessment (MindCrowd) of paired-associate learning (PAL). In a sample of 67 individuals between 18-93 years of age who were imaged at their home or convenient nearby location, we show expected trends in brain volumes with age and detail associations between learning and memory-related brain region volumes and PAL performance. Results demonstrate the ability to collect reliable neuroimaging and cognitive data outside of traditional imaging research settings with important implications for engaging traditionally underrepresented communities in neuroimaging research.

**HIGHLIGHTS:**

1. First demonstration of portable neuroimaging with web-based neurocognitive assessments for routine remote assessment of brain changes associated with aging and age-related cognitive changes.
2. Replication of general brain changes with age and associations with associative learning at low-field strength (64mT) as previously reported at higher routine 1.5T and 3T field strengths.
3. Results demonstrate the ability to collect reliable remote neuroimaging and cognitive test data with important implications for engaging traditionally underrepresented communities in neuroimaging research.

## INTRODUCTION

Neuroimaging has provided salient information on the changing brain tissue macro and microstructure, cortical and sub-cortical morphology and morphometry, and functional connectivity across the lifespan. Studies of volumetric change describe a non-linear pattern characterized by rapid growth of the brain’s white and gray matter throughout infancy and childhood, peaking in the second to fourth decades of life, followed by a slow but progressive decline throughout adulthood [1-8]. Tissue-wise and regional differences exist, with cortical gray matter reaching its maximal value during adolescence while global white matter volume is maximal between 30 and 40 years of age [2, 6, 8]. Subcortical structures, similarly, follow differential growth trajectories, with peak values occurring throughout the second decade of life [5, 7]. Overall, patterns are generally preserved between males and females [8, 9], though absolute volume is, on average, greater in males in large part due to their larger physical body and head size.

Patterns of brain change across the lifespan have further been associated with emerging and receding cognitive skills and abilities. Volume reductions in memory-related regions (e.g., hippocampus) have been associated with age-related memory changes among otherwise healthy older adults, as well as in mild cognitive impairment and Alzheimer’s disease [10-13]. More generally and beyond memory function, global and regional brain volumes have been associated with differential executive functioning skills [12, 14, 15], processing speed [16, 17], and general intelligence [18, 19].

Despite the utility of neuroimaging to the study of healthy aging and neurodegenerative disorders, MRI studies are expensive and often limited to specialized university imaging centers or larger research hospitals. As consequence, they typically have relatively small sample study sizes (n < 30) and rely on populations of convenience, i.e., geographically proximal participants who are able to travel independently or have nearby family members or other support. These factors can bias the study population towards particular sociodemographic phenotypes (e.g., higher socioeconomic and/ or educational backgrounds, individuals living independently with lower disease burden, etc.) that may affect the generalizability of findings and conclusions. Large-scale neuroimaging initiatives such as the Alzheimer’s Disease Neuroimaging Initiative (ADNI) [20], the Human and Lifespan Connectome Projects [21, 22], and the UK Biobank [23] aim to provide study sizes large enough to avoid potential biases (or enable direct modeling of them). These large studies, however, are financially expensive and logistically complex, and still require participating individuals to travel to centralized imaging and research centers.

Over the past 5-10 years, internet and tabled-based tools have made scalable and remote cognitive assessments feasible [24-26]. This trend toward remote assessment has been further accelerated by the COVID-19 pandemic, which forced many research and healthcare centers to seek reliable and reproducible online alternatives to traditional in-person visits and assessments [25, 27, 28]. These tools offer the potential to reach beyond the traditional study populations and include participants from a wide range of geographic, demographic, and socioeconomic backgrounds. MindCrowd [29-32] is one such accessible and easy-to-use web-based platform for cognitive and demographic assessment, designed specifically to overcome challenges with small sample-size studies and to increase inclusive and diverse participation. Participants over 18 years of age are able to anonymously provide general background sociodemographic information (e.g., age, biological sex, education attainment, spoken and written languages, and country of residence). If willing, more granular data may also be provided, including details of medical, health, and lifestyle factors (e.g., marital status, handedness, race, ethnicity, number of daily prescription medications, a first-degree family history of dementia, and yes/no responses to the following: seizures, dizzy spells, loss of consciousness for more than 10 min, high blood pressure, smoking, diabetes, heart disease, cancer, stroke, alcohol/drug abuse, brain disease and/or memory problems). Participants can also optionally provide identifiable name and residential address information for follow-up studies, as well as indicate a willingness to provide biosample collections and participate in ancillary studies.

Cognitive assessment on MindCrowd consists of a simple visual reaction time (svRT) and a paired-associates learning (PAL) task. svRT and PAL are quick and sensitive tests of processing speed and associative episodic memory function, respectively. Past work has shown these cognitive functions are affected in the earliest stages of cognitive impairment and Alzheimer’s Disease (AD), but also reflect the general decline in cognitive performance in healthy aging. The ability to capture data from hundreds of thousands of participants at a relatively low cost and without time-consuming and intensive in-person visits has allowed the team behind MindCrowd to investigate the impact of diverse family, medical history, and genetic factors on cognitive performance across the adult lifespan [29, 32], and to identify potential cases of previously undiagnosed cases of cognitive impairment and dementia [31].

While reliable remote and internet-based cognitive assessments are becoming increasingly common, remote collection of MRI data has been impracticable due to the size and weight of common 1.5 and 3 Tesla (T) systems, as well as their electrical requirements, and helium and maintenance needs. Though semi-trailer 18-wheeler mounted 1.5T systems are broadly available throughout North America, Europe, and Asia, they share the size, weight, and electrical needs of their sited brethren, and are designed for institutional use as adjuncts to static systems installed at hospitals or clinics, or as semi-permeant solutions for smaller institutions. These systems require specially installed concrete parking pads and high voltage electrical supplies and are not designed for use at a participant’s home. However, advancements in MRI systems that operate at lower magnetic field strength (I.e., less than 100mT) with permanent or resistive magnet arrays present an alternative to conventional systems with the possibility of enabling “residential MRI” - truly remote neuroimaging performed at a participant’s home, assisted living facility, or other residential or convenient and nearby location (e.g., library, shopping center, etc.) [33]. While lower field systems are not replacements for higher field strength scanners, and currently offer a limited repertoire of imaging contrasts and methods, they do allow for high-quality anatomical imaging [34] and have shown replication of developmental patterns observed at higher field strength [35].

As much of the past work with low field scanners has focused on clinical applications (e.g., identification of pediatric hydrocephalus [36], multiple sclerosis [37], stroke [38], and other indications [39]), its utility in neuroscience research, such as associations between brain morphology and cognitive performance, remains unknown. Like remote cognitive assessments, the ability to reliably collect high-quality and information-rich MRI data at a participant’s home could bring new opportunities to the study of aging, cognitive decline, cognitive impairment, and dementia. Remote MRI would allow the inclusion of participants with mobility challenges or who lack transportation options, those who live long distances away from research centers and outside traditional recruitment areas, and those with competing family, work, or other time commitments. The reduced expense of low field strength MRI may further allow for increased study population size, improving statistical power and generalizability of study findings.

To this end, in this study we sought to determine the feasibility of combining remote cognitive assessment via MindCrowd with mobile low field MRI in a modified Ford Transit cargo “scan van” equipped with a 64mT MRI scanner [40] to (1) Determine the feasibility of collecting reliable remote MRI and cognitive data in adults and elderly individuals; and (2) Replicate previously reported associations between regional brain volumes and cognitive performance with an established cognitive assessment, PAL, with substantial normative data against which to compare.

## METHODS

This study was performed in accordance with ethics approval and oversight by WCG IRB and the Rhode Island Hospital Institutional Review Board. All volunteers provided informed consent for both the MRI and neurocognitive assessment components.

### Participants

A total of 75 individuals (42 female, 56%) between 18 and 94 years of age were recruited to participate in this study. Of these, 67 (39 female) completed the web-based cognitive assessments within 1 week of scanning and had high-quality MRI. 8 individuals did not complete the MindCrowd assessments in a timely fashion and/or had poor quality or incomplete MRI (i.e., were positioned too low in the coil). Participant demographics are provided in **Table 1**. The mean age of the final study population was 54.2 ± 19.7 years. In addition to providing age and biological sex, participants were also asked to indicate their education attainment on a 5-item scale (1=Some High School, 2=High School Diploma, 3=Some College, 4=College Degree, and 5=Graduate Degree).

**Table 1.**
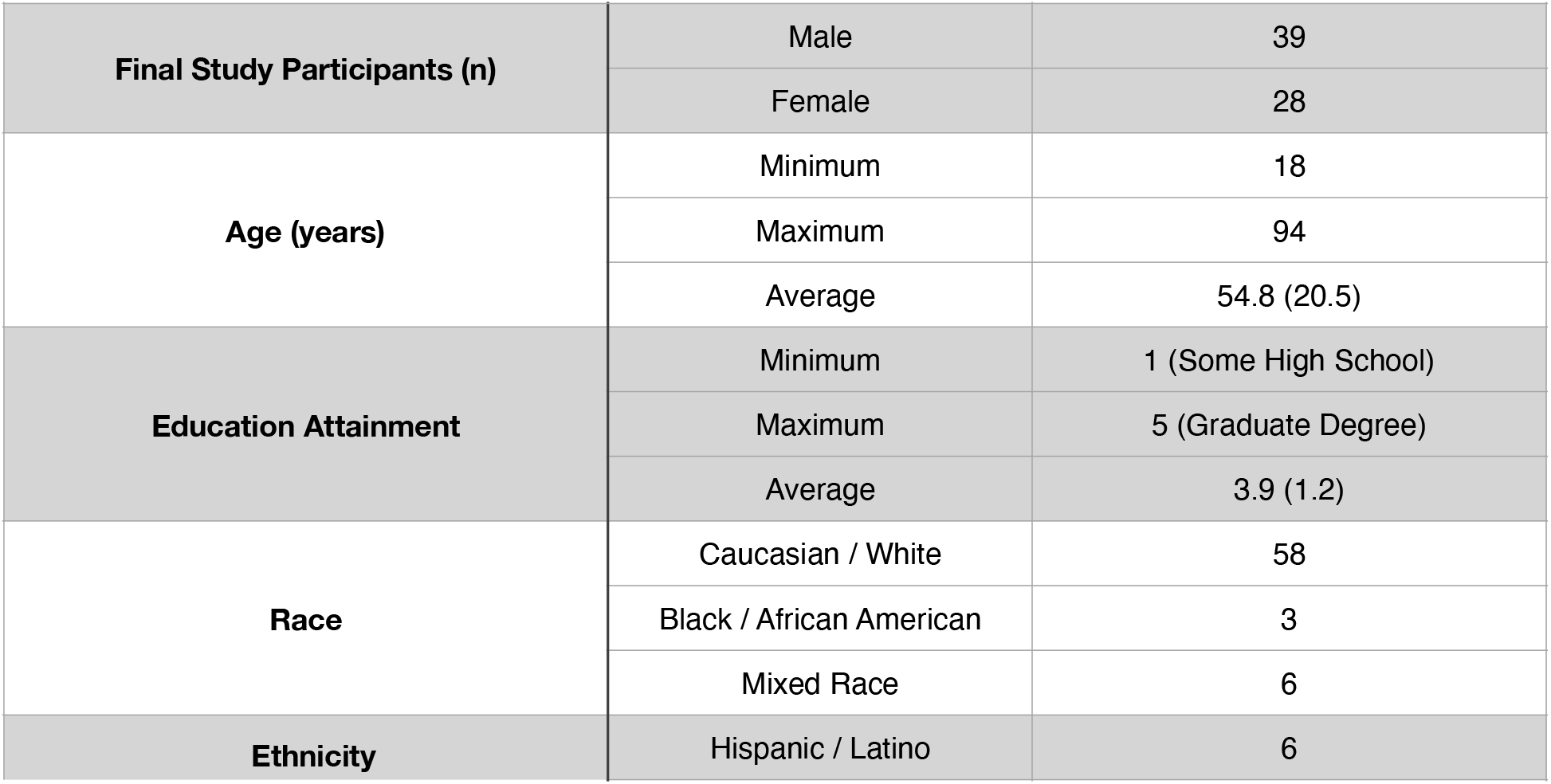
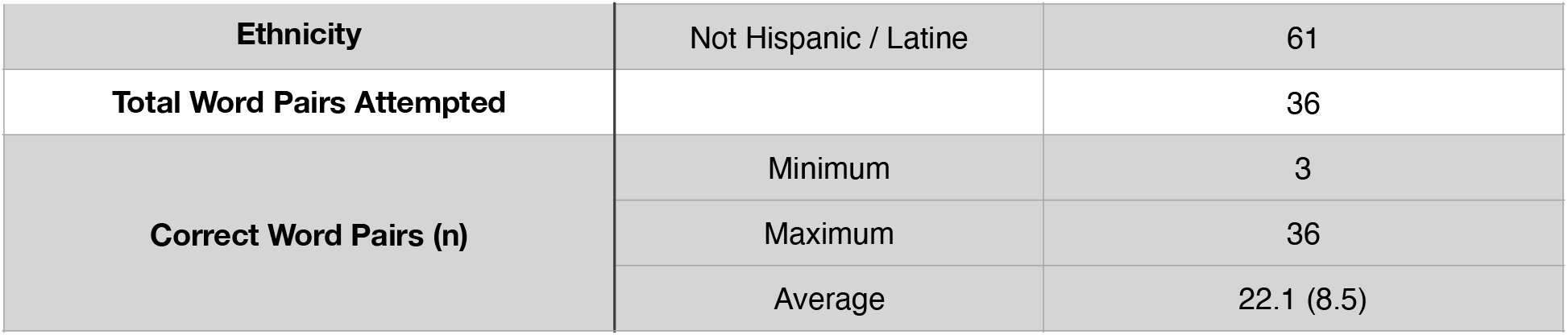
General demographic details and summary cognitive measure statistics for the final study cohort.

Recruitment, MRI scanning, and online assessment collection for this study were performed over a 3-week span. Scanning was performed either at the participant’s home or at a community center ‘pop-up’ event. Total imaging time, including set-up, was ∼20 minutes per subject. Set-up time was slightly longer for individuals who required help into the van and onto the scanner bed. At the pop-up events, 2-3 scans were performed per hour with 8-12 individuals consented and scanned over a ∼6-hour afternoon. Overall, the scanning process was well-tolerated by participants and their families. Family members were able to be in the van during scanning or could wait outside watching through the van’s sliding door (**Fig. 1**).

**Figure 1.**
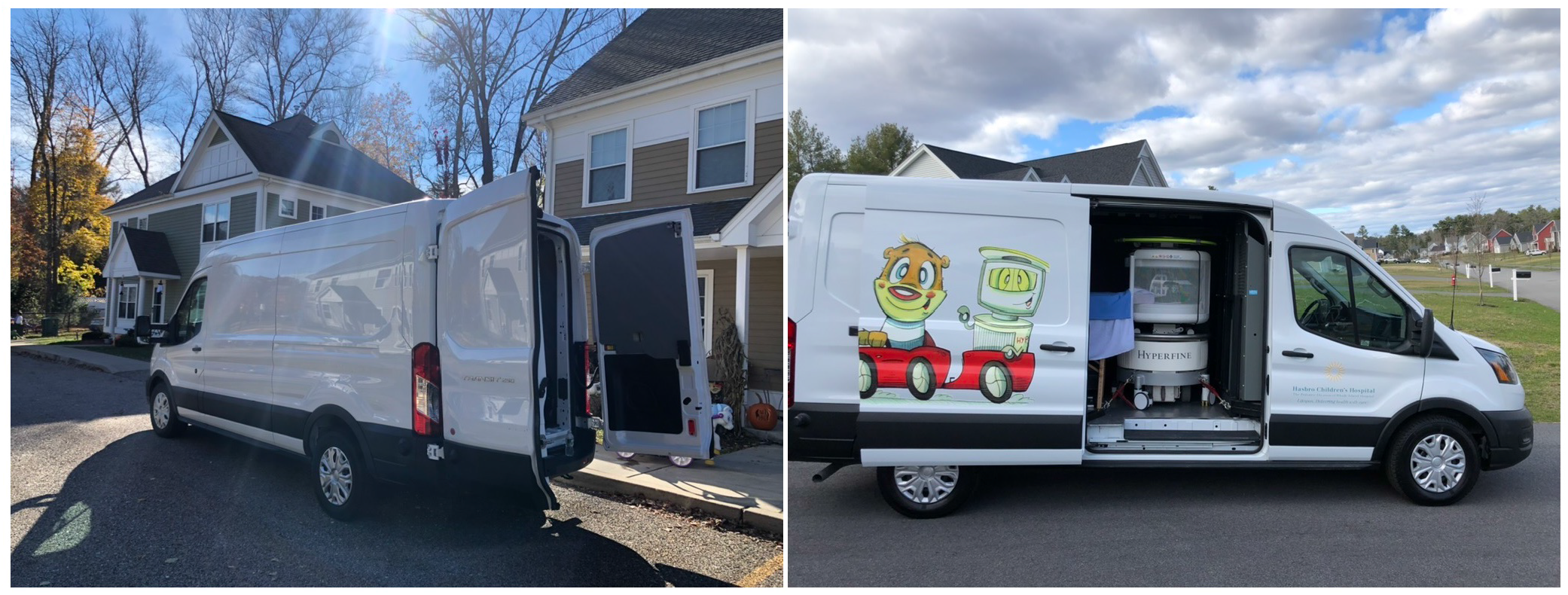
Example photos of the Scan-Van at participant homes. Participants enter the van from the rear doors. The side door can be opened to reduce claustrophobia, allow extra cooling, or just improve the general participant experience.

### Neuroimaging

MRI was performed on a 64mT Hyperfine Swoop system installed in a purpose-modified Ford Transit cargo “scan van” [41] at the participant’s residence or nearby convenient location (e.g., **Fig. 1**). A series of anisotropic resolution T_2_-weighted 3-D fast spin echo (FSE) images were acquired in each of the three orthogonal orientations (axial, sagittal, and coronal) with an in-plane spatial resolution of 1.5mm x 1.5mm and a slice thickness of 5mm. While T_1_-weighted contrast is the standard for anatomical studies at 1.5T and 3T, we have found the reduced T_1_ at 64mT reduces the quality of the T_1_-weighted images, whereas T_2_ provides improved and more consistent tissue contrast across the age span.

A super-resolution reconstruction approach [34] was used to combine the orthogonal images into a single (1.5 × 1.5 × 1.5)mm^3^ image for follow-on processing.

A general study template was constructed using the ANTs 2.2 [41], antsMultivariateTemplateConstruction2.sh script, and the non-linear transformation matrix between this template and MNI space was calculated. The non-linear transformations and the corresponding Jacobean matrices, between each individual’s image and the study template, were also calculated using ANTs.

### Cognitive Assessments

Following neuroimaging, participants were provided with login details and instructions to perform the PAL memory test on MindCrowd (mindcrowd.org). For those without personal internet access, a MacBook Pro laptop or cellular internet-connected tablet was made available for them to use.

For the online implementation of the PAL task, participants are provided an initial learning phase in which they are presented with 12 word-pairs, one word-pair at a time (2 sec. per word-pair). Pairs include 8 difficult associations (e.g., apple-pen) and 4 easier associations (e.g., black-night). Immediately following the presentation of pairs, participants are presented with the first word of each pair and are asked to use their keyboard to type (i.e., recall) the missing word. This learning-recall procedure is repeated for two additional trials. Prior to beginning the task, each participant receives a practice trial consisting of 3 word-pairs not contained in the 12 used during the test. Word-pairs are presented in different random orders during each learning and each recall phase. The same word pairs and orders of presentation are used for all participants. The dependent variable/criterion used in this study was the total number of correct word pairs entered across the three trials (i.e., 12 × 3 = 36, a perfect score).

### Analysis

Following data collection, an initial exploratory voxel-based analysis was performed to identify potential associations between the collected cognitive measures (word-pair association score and reaction time) and brain gray matter density. Following nonlinear alignment to the study template, correction for the effects of the warping on the voxel-wise density measures, and subtle blurring with a 4mm Gaussian kernel, a general linear model was fit at each voxel that modeled PAL as a function of local gray matter density (GM), subject age, and biological sex, i.e.,

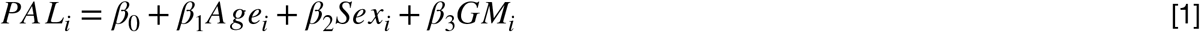

Recognizing prior demonstrated associations between education level and metrics of brain morphometry in healthy and cognitively impaired individuals [42, 43], we extended our exploratory analyses to include reported education level as an additional model variable,

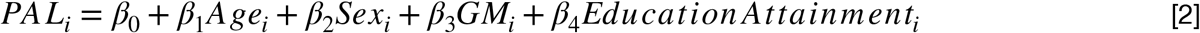

Education was scored on a 5-point scale ranging from 1 (high school diploma) to 5 (graduate or professional degree).

Analysis was performed using the Randomise tool of the FMRIB Software Library (FSL) [44]. Threshold-free cluster enhancement (TFCE) [45] was used to control for the multiple voxel-wise comparisons.

Based on the outcomes of our exploratory analyses, hypothesis-based analysis was performed in which we examined associations between deep brain gray matter structure volumes (including right and left hemisphere hippocampus, amygdala, caudate nucleus, thalamus, globus palliudus, and putamen) and PAL scores. To calculate the deep gray matter structure volumes, the Oxford-Harvard subcortical atlas provided as part of FSL and registered to MNI space [46] was aligned to each participant’s image using the inverse of their individual->study template and study template->MNI template transformations. The aligned structure masks were then thresholded at 0.95 and the volumes of the inscribed regions were calculated.

In addition to sub-cortical gray matter volumes, we also used this atlas approach to examine the associations between total brain white matter (WM), gray matter (GM), and cerebral spinal fluid (CSF) volume and PAL scores. For these tissue volumes, the MNI structural atlas was used as the reference [47, 48].

We hypothesized that brain regions involved in memory and learning networks, i.e., hippocampus, amygdala, caudate nucleus, and thalamus would be significantly associated with word-pair scores [49-55]. This hypothesis was tested using a series of linear models that included PAL score and structure volume, as well as subject age, biological sex, and total brain tissue volume (the summation of white and gray matter volumes),

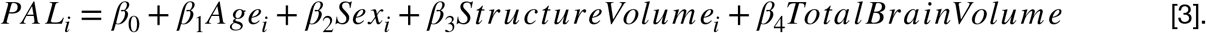

The Holm-Bonferroni approach was used to account for the 15 independent tests.

A simple model omitting the Structural Volume term was also fit to the data, allowing us to determine the additional variance explained by this variable.

## RESULTS

As a first check of data consistency, we plotted the total number of correct word pairs against subject age (**Fig. 2**), showing a decrease in performance as a function of age in agreement with past findings showing a decrease of 1-2 word-pairs per decade [29, 56].

**Figure 2.**
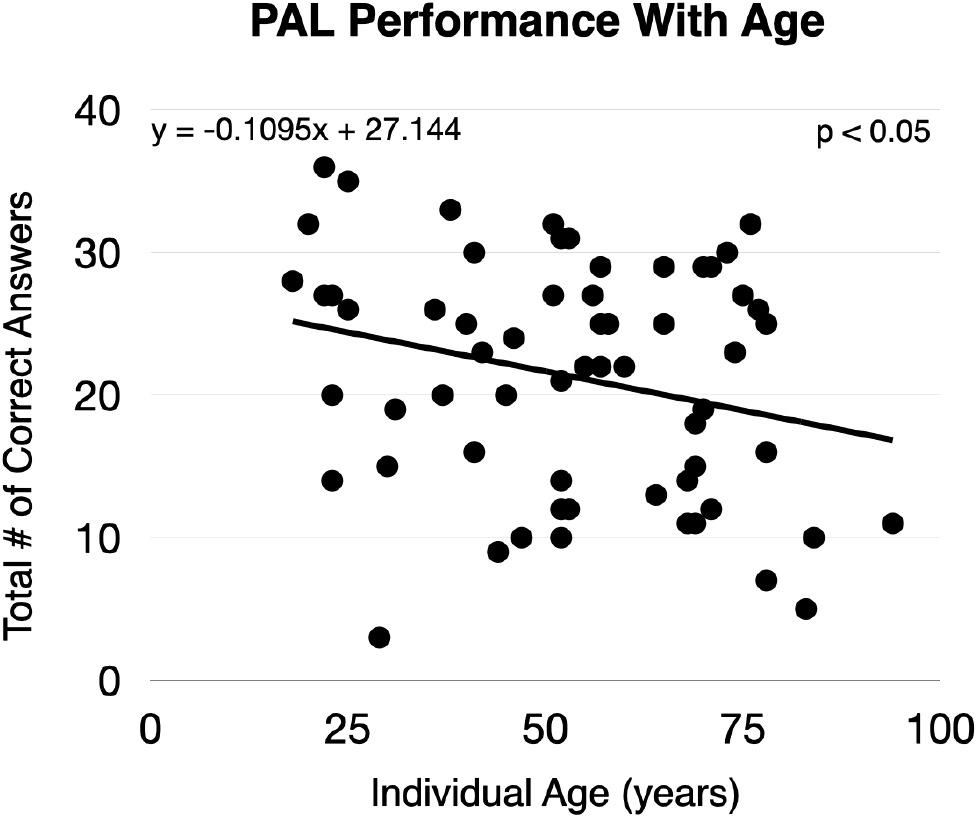
Number of correct word pairs as a function or participant age.

To examine the MRI data, we plotted total brain white and gray matter volume percentage versus age (**Fig. 3**), observing the expected quadratic and linear trends for white and gray matter, respectively.

**Figure 3.**
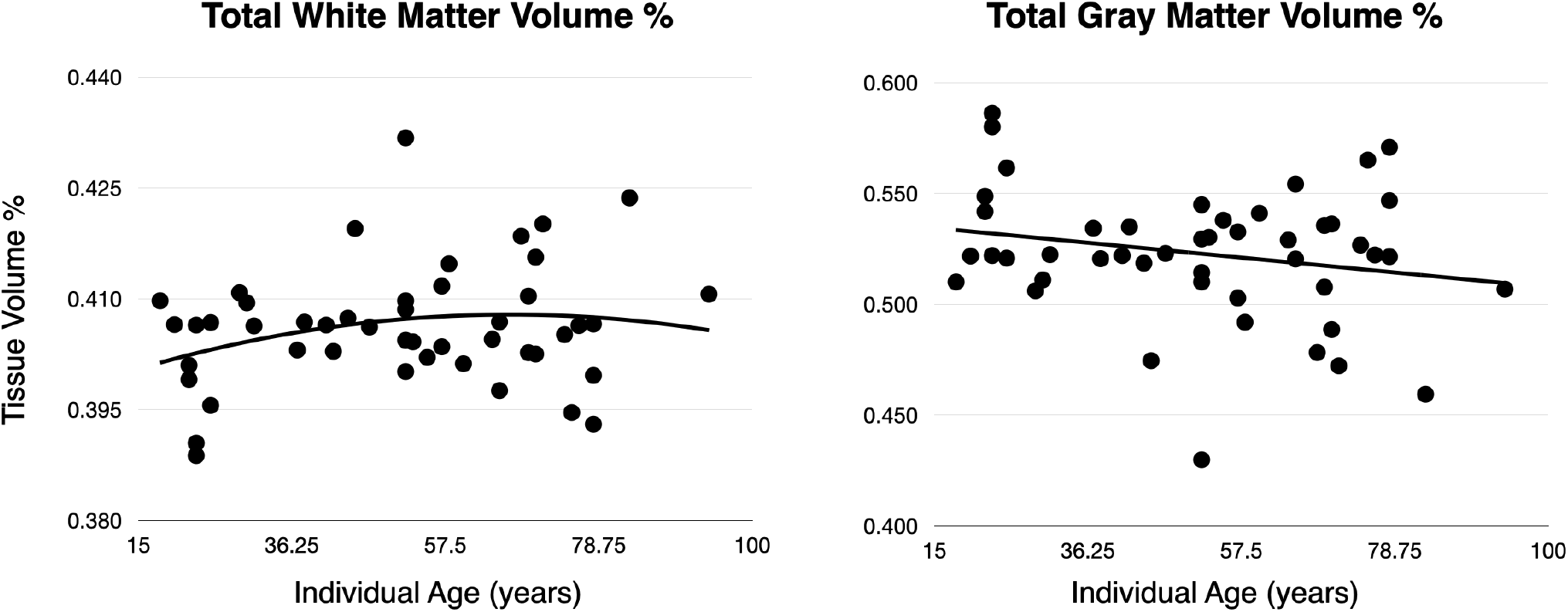
Representative plots of percent white and gray matter volume as a function of individual age.

Figure 4. contains the results of our exploratory voxel-based analyses, highlighting brain areas with significant (p<0.05 FWE) associations between gray matter density and PAL test scores. Modeling the association between local gray matter density and PAL performance, controlling for participant age, biological sex, and education attainment, we found significant associations predominately in left hemisphere regions, including hippocampus, parahippocampal gyrus, inferior temporal gyrus, thalamus, putamen, frontal pole and orbital cortex, caudate, and Broca’s area. In addition, right hemisphere precentral gyrus was also identified. These results did not substantively differ from the model that only included age and biological sex as additional variables of non-interest.

**Figure 4.**
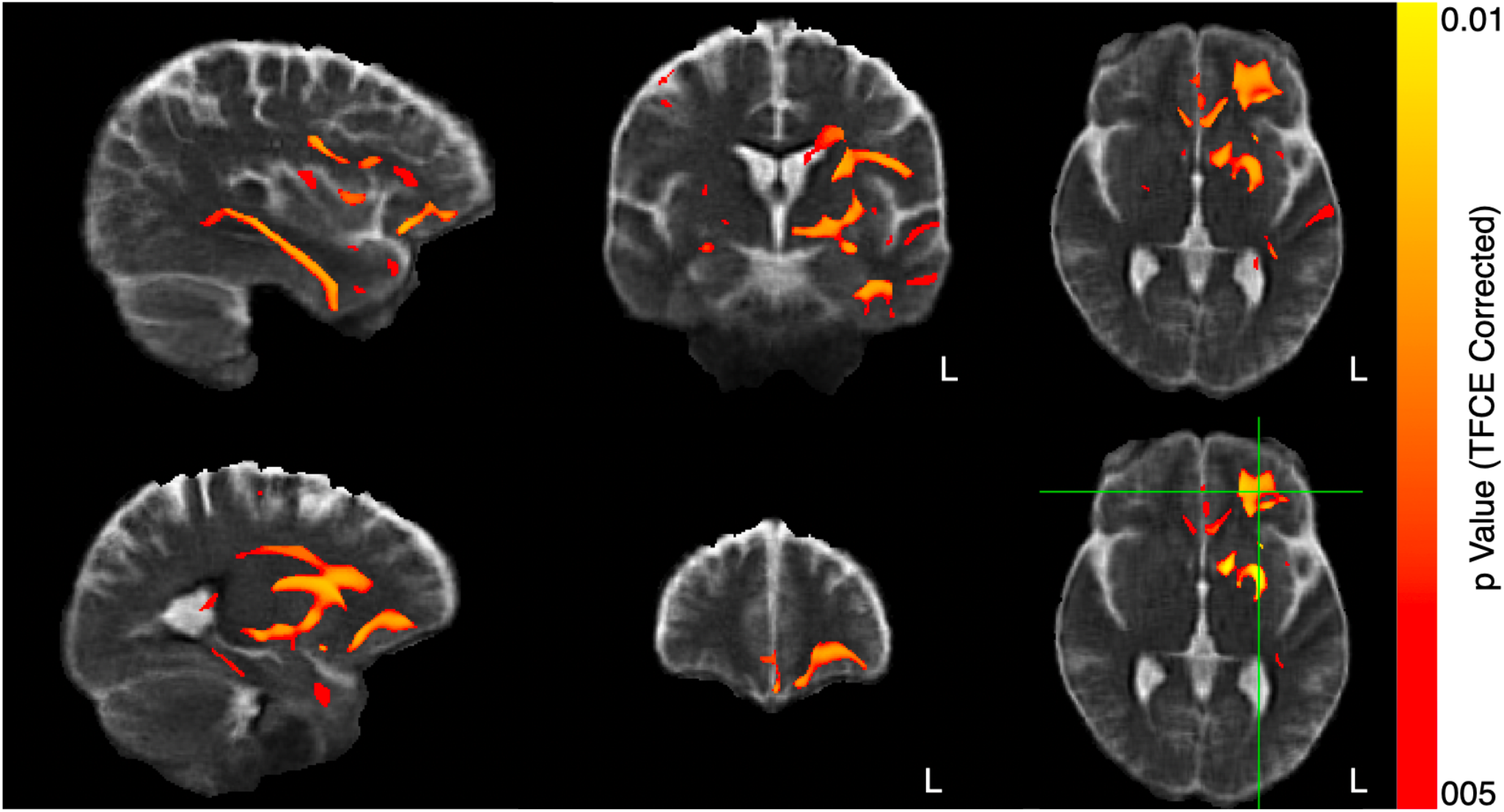
Exploratory voxel-based morphometry analysis examining associations between gray matter density and PAL score, controlling for subject age, biological sex, and education attainment. Highlighted regions denote significant associations (corrected for multiple comparisons using threshold-free cluster enhancement).

**Table 2** contains the results of the linear modeling of regional subcortical gray matter volumes and PAL test scores, with representative plots of predicted vs. actual test scores as a function of regional volume, which are shown in **Figs. 6** and **7**.

**Table 2.** Results of our general linear model analyses showed significant (p < 0.05 corrected associations between individual brain tissue and subcortical gray matter structure volumes and assessed PAL performance controlling for age, biological sex, and total brain volume. Measures in bold denote significant associations after correction for multiple comparisons.

| Brain Region | Coefficient | Unadjusted p Value | Corrected p Value | Additional Explained Variance |
| --- | --- | --- | --- | --- |
| Whole Brain WM | 0.0012 | 0.0105 | 0.0197 | 0.1250 |
| Whole Brain GM | 0.0006 | 0.0067 | 0.0168 | 0.1250 |
| CSF | <0.0001 | 0.9772 | 0.9772 | 0.0000 |
| Left Amygdala | 0.0749 | 0.0004 | 0.0054 | 0.2053 |
| Right Amygdala | 0.0713 | 0.0120 | 0.0199 | 0.1087 |
| Left Caudate | 0.0170 | 0.0059 | 0.0168 | 0.1287 |
| Right Caudate | **0.0268** | **0.0135** | **0.0203** | **0.1052** |
| Left Hippocampus | **0.0225** | **0.0053** | **0.0168** | **0.1318** |
| Right Hippocampus | **0.0284** | **0.0061** | **0.0168** | **0.1278** |
| Left Pallidum | 0.0160 | 0.2235 | 0.2394 | 0.0268 |
| Right Pallidum | 0.0307 | 0.0770 | 0.0889 | 0.0751 |
| Left Putamen | 0.0096 | 0.0398 | 0.0543 | 0.0903 |
| Right Putamen | 0.0127 | 0.0465 | 0.0582 | 0.0874 |
| Left Thalamus | **0.0052** | **0.0023** | **0.0168** | **0.1547** |
| Right Thalamus | **0.0049** | **0.0089** | **0.0191** | **0.1170** |

Results of the regional analyses build on the exploratory outcomes, showing hippocampus, amygdala, caudate, and thalamic volumes, as well as whole-brain white and gray matter, were significant predictors of PAL performance after correction for multiple comparisons (**Table 2**). In addition, right and left hemisphere putamen showed a trend towards significance. In each of these cases, the addition of the regional volume measure explained an additional 10 to 20% of the total variance in PAL performance. These numerical results are graphically represented in **Figs. 5** and **6**, which show (Fig. 6) the measured PAL scores for each individual and predicted PAL by the linear model based on regional volumes as a function of age. The relationship between the measured and predicted PAL scores is shown in **Fig. 6**. As expected, we observe stronger correlations for brain regions where regional volume was a significant predictor of performance (e.g., white and gray matter vs. CSF volume).

**Figure 5.**
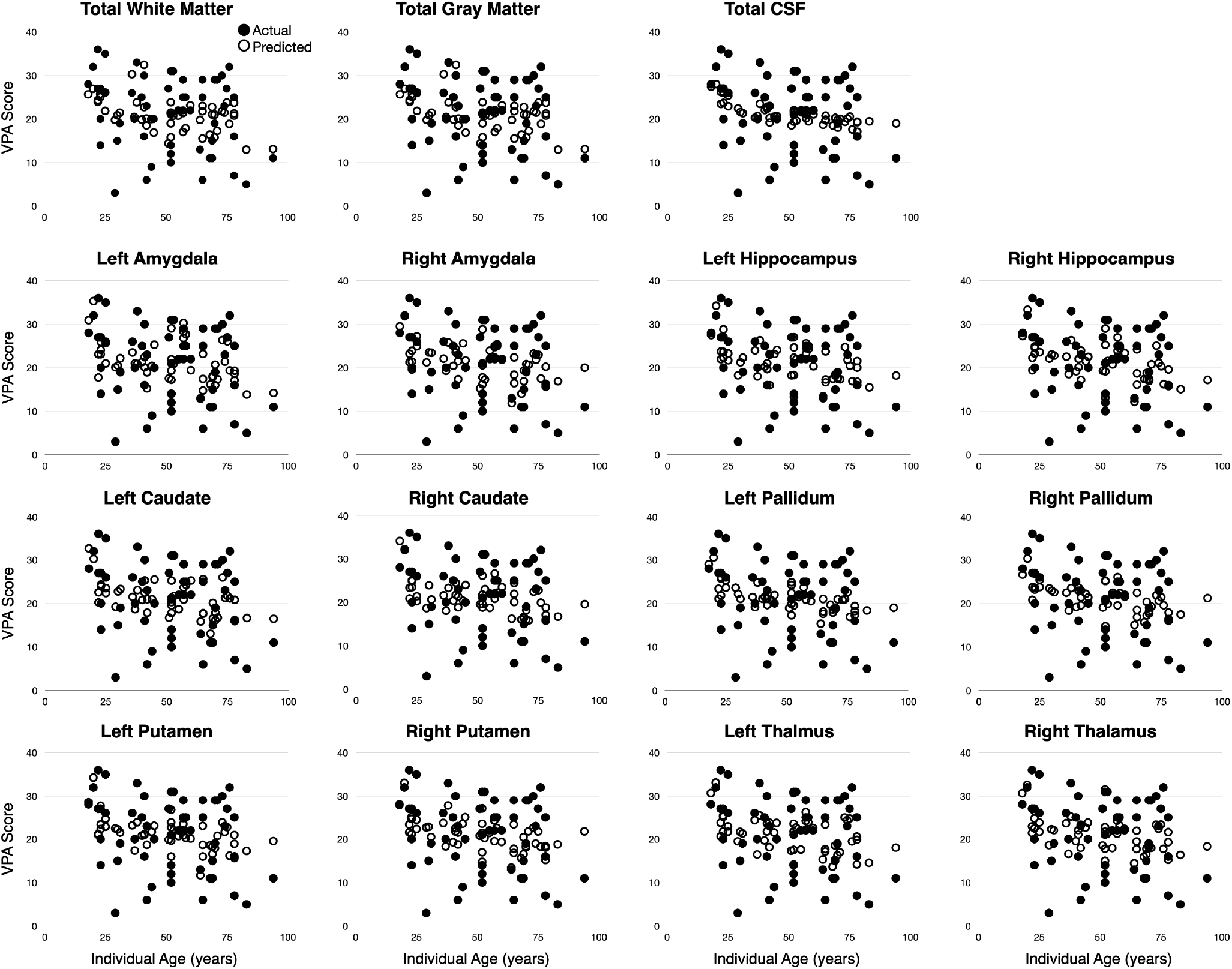
Results of the regional volume analysis. For each region, we plot the measured PAL scored vs. age as well as the predicted PAL score calculated from the estimated regression model.

**Figure 6.**
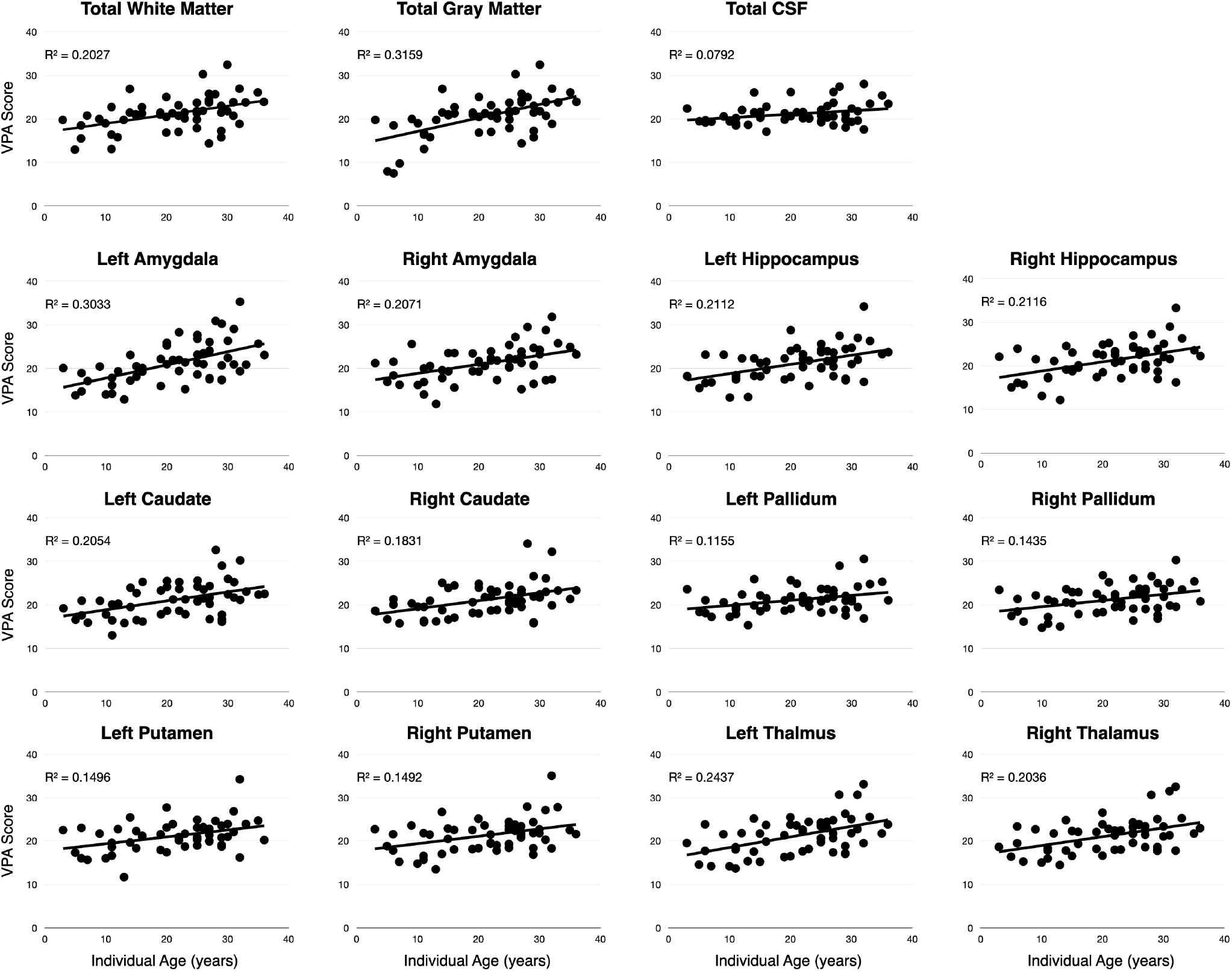
Results of the regional volume analysis. For each region, we plot the measured PAL scored vs. the predicted PAL score calculated from the estimated regression model.

## DISCUSSION

Here we have presented the first preliminary reports examining the practical use of a van-based portable low-field MRI scanner and web-based cognitive assessment to investigate changing total and regional brain volumes associated with cognitive performance. Results agree with prior reported outcomes, replicating the established general trends of white and gray matter change with age and identifying significant brain volume associations with memory performance in previously reported subcortical gray matter volumes. From our exploratory analyses, we found significant positive associations between improved PAL performance and increased local gray matter density in left hemisphere deep brain regions (hippocampus, thalamus, caudate, and putamen) as well as the left parahippocampal gyrus, frontal pole and Broca’s area (Brodmann area 44), and right precentral gyrus (Brodmann area 4). These regions have known functional associations with language processing, memory, and learning. While the precentral gyrus is not directly related to these neurological functions, it is involved in movement and may reflect individual differences in tablet use or ability during the MindCrowd assessment. These results were confirmed in follow-up regional analysis, which identified significant brain-PAL associations in bilateral hippocampus, amygdala, thalamus, and caudate, controlling for subject age, sex, educational attainment, and total brain volume. Although the putamen associations did not survive FWE correction (with corrected p values of approximately ≃ 0.05) they do show a strong trend towards significance.

These results carry important implications for future opportunities and directions in neuroscience research. The combination of mobile at-home neuroimaging and web-based remote assessment presents an important new opportunity to engage individuals from socially, economically, and educationally disadvantaged communities that are often under-represented in clinical and public health research [58, 59]. Moreover, some clinical and pre-clinical populations (e.g., individuals in their 30s or 40s with a family history of Alzheimer’s Disease but without personal memory or cognitive complaints) are often difficult to recruit and/or retain in long-term longitudinal trials and studies because of family and work time commitments. By bringing the scanner to them at home or work, and allowing cognitive assessments to be performed on their time, involving these important cohorts may be less challenging. Though our study sample were predominately from higher educational (∼34% reported a college or post-graduate degree) and non-Latino/Hispanic Caucasian (86%) backgrounds, there is nothing that fundamentally limits our approach from reaching broader individuals and communities.

While memory changes are experienced by most, but not all, individuals as they age, worsened associative memory performance can also be indicative of emerging cognitive impairment and dementia [55, 57-59]. Neuroanatomical correlates of reduced performance on various episodic memory tests, including associative memory, include reduced total gray matter volume and regional reductions in the hippocampus, thalamus, and putamen. Alongside these functionally-related changes, progressive volume loss in entorhinal cortex, frontal lobe, and temporoparietal cortical areas have been implicated in mild cognitive impairment (MCI) [60]. Increased rates of volume loss in these regions are potentially predictive of progression from MCI to Alzheimer’s Disease [61].

A recent report on the state of study needs in aging research [62] has estimated that nearly 100,000 participants will need to be recruited into the existing set of US-based observational and clinical trials for prospective preventative or therapeutic AD treatments. The authors further estimate that achieving this level of enrollment will require screening upwards of 1 million potential participants and their families. As with other scientific and clinical health research, research in aging and AD has suffered from under-representation of individuals from racial and ethnic minorities, and economically disadvantaged communities [63] - despite these groups having a higher potential risk for dementia [64]. A further missing gap is individuals who are free of clinical symptoms but at risk for AD, since initiating pathology may appear 2 or 3 decades before overt memory loss or other symptoms become apparent [65]. This necessitates recruitment and longitudinal retention of 30 to 50-year-old individuals. These individuals, however, often have busy family, work, and social schedules, which inhibits participation in research studies. To address these challenges, Watson et al. [62] highlight the need to consider and accommodate the location and time needs of participants and their families (or study partners) by conveniently locating study sites or, ideally, performing study visits at the participant’s home.

The results presented here highlight the potential to perform participant screening, enrollment, and at-home study visits, even in the context of neuroimaging studies. While we have presented MindCrowd as an effective tool for remote cognitive assessment, it can also be envisaged and utilized as a screening tool, allowing the identification of individuals willing to participate in research studies, and with important clinical and/or sociodemographic phenotypes. Beyond cognitive and health and family history information, participants can also provide biological samples from at-home collections (e.g., saliva samples for genomic analyses) allowing genetic phenotypes to also be screened. We purposefully designed our Scan-Van on a 2021 Ford Transit Van base (high roof and extended length 2500 model with a 9500lbs gross vehicle weight rating) to achieve three functional aims: 1. Ability to travel on local and dirt roads to allow access to rural communities and not rely on truck or high weight capacity routes. 2. To be driven by anyone with a regular driver’s license (i.e., not require a commercial CDL license). And 3. Have access to a large national network of maintenance and repair facilities with ready access to parts and service. In addition, we also designed the system to use an EGO Power+ 3000W portable power station that provides more than 6hrs of continuous scanning from 4 rechargeable (with additional batteries able to be hot swapped to allow longer scanning), and for the ability to load and unload the scanner for imaging in or outside the vehicle.

While 5 scans were rejected due to poor subject positioning in the coil (all collected on the same day of scanning), this error was corrected and no further scans were rejected for this or other quality reasons (e.g., motion artifacts, noise, poor contrast, etc.). As all scans for this study were performed in the van, which required participants to be mobile enough to walk up 2 steps (∼18 inches) into the van and then onto a 30” high massage bed. For larger-scale studies, the scanner can be removed from the van to accommodate participants with mobility challenges and unable to get into the van. To allow the scanner to be used in the fall and winter months (in New England) and avoid participant discomfort or operating outside of the scanner’s recommended range (5-30C), a heating system was built into the van that could be complemented with a portable electric heater (also run from the portable battery station). In the summer months, operating with the rear doors open and an oscillating fan provides sufficient comfort for the short scan duration without impacting scan quality. In cases of extreme heat, a roof-mounted air-conditioning unit can also be used.

While most prior studies in aging and AD, including the original ADNI protocol [66], focused on structural and morphology changes, more recent investigations have included assessment of tissue microstructure (diffusion tensor imaging, DTI), cerebral perfusion, and structural and functional connectivity. Currently, the Hyperfine system is capable of four structural image contrasts (T_1_, T_2_, T_2_-FLAIR, and single-axis DWI). As more research groups gain access to these portable and lower field strength systems, it is likely we will see steady improvements in acquisition techniques, including DTI, relaxometry, and potentially perfusion imaging.

A potential source of variability that was not directly accounted for here is the type and size of the device used for the MindCrowd assessment. Participants were provided either a 13” MacBook Pro or a 10.9” iPad Air to complete the online cognitive assessments at the van. Participants who completed the assessments at home may have used a differently sized laptop/desktop computer, tablet, or mobile phone. These differences in screen size and keyboard size and type (physical vs. virtual touch screen) may have impacted PAL performance and added additional variability to PAL performance. While further investigation is needed to understand this potential variability (and accurately account for it), we do not believe it will substantively alter the presented results. For the PAL assessment, individuals are allowed 10s to begin their response and are allowed ample completion time before the system moves on to the next word-pair. Thus, challenges with a smaller or unfamiliar keyboard should not significantly reduce their total score.

A general challenge with online unassisted assessment is knowing when low scores are accurate reflections of an individual’s ability or are the result of them not paying attention during the assessment or not understanding/following instructions. For example, in our cohort, 4 individuals under 45 years of age reported PAL scores less than 10. These scores are more than 3 standard deviations below the expected mean for healthy individuals of their age [32]. As an ad-hoc analysis, we repeated our regional volume analyses excluding these individuals as outliers. The results of this analysis did not differ from those presented in **Table 2**, with the exception that right putamen volume remained a significant predictor after Holm-Bonferroni correction for the multiple tests.

In summary, the work described here demonstrates the feasibility of coupling mobile neuroimaging and web-based cognitive assessments. With this approach, we replicate known effects of aging on associative memory performance as well as highlight neuroimaging-based changes in learning and memory-based regions of the brain with performance. We propose that the combination of a mobile neuroimaging laboratory with on-demand web-based cognitive assessment has significant potential for the future study of many types of disadvantaged and understudied populations, including race-ethnic, socioeconomic, time-constrained, and geographically-distanced groups. More work is necessary to refine our approach and appropriately tailor recruiting and retention practices to ensure success with such groups, however, we propose that our work demonstrates the significant potential for these efforts.

## Data Availability

All deidentified data is avaialble upon request.

## ACKNOWLEDGEMENTS

The authors extend important thanks to all study volunteers and their families, as well as Neil Sharpe and the Brain Injury Association of Rhode Island.

